# Choice of estimands and estimators affected the interpretation of results for some outcomes in a cluster-randomised trial (RESTORE) due to informative cluster size

**DOI:** 10.64898/2026.05.05.26352371

**Authors:** Dongquan Bi, Andrew Copas, Fan Li, Michael O Harhay, Brennan C Kahan

## Abstract

**Background and objective:** In cluster-randomised trials (CRTs), different estimands can be targeted, such as the individual-or cluster-average effect. These two estimands can differ in magnitude when outcomes or treatment effects vary with cluster size (termed informative cluster size). When informative cluster size is present, commonly used estimators for CRTs, such as mixed-effects model and generalised estimating equations with an exchangeable correlation structure (termed GEEs(exch)), can be biased for both these estimands. With little documented evaluation of When informative cluster size, it is currently unknown how commonly it occurs in practice. The aim of this work was to explore whether informative cluster size is present in a published CRT and to investigate its impact on trial results.

**Methods:** We re-analysed the RESTORE CRT, which compared protocolised sedation with usual care for critically ill children. For each outcome, we first modelled the association between cluster size and outcome/treatment effect; next, we assessed the impact of informative cluster size by comparing differences between (i) individual-vs. cluster-average estimates and (ii) estimates from mixed-effects models and GEEs(exch) (which can be affected by informative cluster size) to those from IEEs (which are robust to informative cluster size).

**Results:** We found evidence of an association between cluster size and either outcomes or treatment effects for 16/33 outcomes (48%). This led to statistically significant differences between the individual- and cluster-average treatment effects for 5 of 33 outcomes (15%). There were >10% differences between (i) individual- and cluster-average treatment effect estimates for 17 outcomes (52%) and (ii) estimates from mixed-effects models/GEEs(exch) and estimates from unweighted IEEs for 13 outcomes (39%). For some outcomes, differences in the choice of estimator or estimand led to differences in the interpretation of results. For example, for the outcome postextubation stridor, the individual-average estimate showed a significant harmful effect (OR=1.65, 95% CI 1.02 to 2.67), unlike the cluster-average (OR=1.38, 95% CI 0.87 to 2.19) or GEEs(exch) estimate (OR=1.57, 95% CI 0.98, 2.50).

**Discussion:** informative cluster size can occur in CRTs, and the use of estimators that are not clearly aligned to the target estimand can affect the interpretation of some results.

**What is new?:** *Key findings:*

- This re-analysis of the RESTORE cluster randomised trial found that choice of estimand and estimator could affect the interpretation of results for some outcomes

*What this adds to what is known:*

- This work provides empirical evidence that informative cluster size can occur in cluster randomised trials, and can affect results based on the choice of estimand or estimator

*What is the implication and what should we change now:*

- Trialists should clearly define their target estimand and choose an estimator that is aligned to that estimand
- Careful consideration of the plausibility of assumptions underpinning each estimator, including the likelihood of informative cluster size, can help ensure appropriate analysis methods are used
- When mixed-effects models or GEEs with an exchangeable correlation structure are used, sensitivity analyses using independence estimating equations or other appropriate methods should be used to evaluate the robustness of results to informative cluster size

## 1. Introduction

Cluster-randomised trials (CRTs) involve randomising groups of individuals (such as hospitals, schools, or communities) to different interventions. [1, 2] Recent studies have shown that different estimands (i.e. treatment effects) can be targeted in CRTs, such as the individual-average or cluster-average effect. [3–5] These differ in how individuals and clusters are weighted in the definition of the estimand. [3, 4] The individual-average estimand assigns equal weight to each individual, while the cluster-average estimand assigns equal weight to each cluster. Both estimands can be appropriate depending on trial objectives. [3]

A key challenge that has emerged is how to ideally estimate these different estimands, as there is growing recognition that different methods of analysis (estimators) can target different estimands and may be biased for some or all of them. [3, 4, 6–10] This can occur when there is informative cluster size (ICS), that is, when outcomes or treatment effects vary with cluster size. Under ICS, the individual- and cluster-average estimands typically differ, and estimators targeting one will be biased for the other. [3, 4] Common estimators in CRTs, such as generalised estimating equations (GEEs) with a working exchangeable correlation structure (termed “GEEs(exch)” hereafter) and mixed-effects models with a random intercept, may be biased for both. [3, 4, 6, 11] This occurs because mixed-effects models and GEEs(exch) weight clusters by their inverse-variance (a function of the intraclass correlation coefficient and the cluster size), which corresponds neither to the weighting used for either the individual-or cluster-average estimand. [3, 11, 12]

An unbiased alternative under ICS is independence estimating equations (IEEs), [3, 4, 8–11, 13–15], which involves specifying a working independence correlation structure with cluster-robust standard errors. [16] However, while IEEs can provide unbiased results, they may be less efficient than mixed-effects models and GEEs(exch). [3, 17] There has been little documented evaluation of ICS; to our knowledge, only one study has systematically evaluated the occurrence of ICS across all outcomes in a CRT, [6] which found that ICS may have occurred. It is, however, difficult to reach definitive conclusions on this basis, as it was a single study in a specific clinical setting and limited to 6 clusters.

Given the scarcity of evidence around the likelihood of ICS in practice, it is difficult for trial investigators to make informed decisions about which estimators to use. If ICS rarely occurs, the efficiency gains from GEEs(exch) and mixed-effect models are likely to outweigh the small chance of bias from ICS. However, if ICS is common, the risk of bias is likely unjustified regardless of the efficiency gains. The purpose of this paper is therefore to systematically re-analyse a published CRT to add further evidence on the likelihood of ICS and to evaluate to what extent it may impact trial results.

## 2. Methods

The purpose of this study is to assess the presence and impact of ICS in the RESTORE trial. To do this, we first modelled the association between cluster size and outcomes/treatment effects to look for the presence of ICS. Next, we evaluated whether ICS could affect results by comparing different estimands and estimators. Below, we describe the RESTORE trial (section 2.1), followed by the methods used to model the association between cluster size and outcomes/treatment effects (section 2.2), and finally the methods used to evaluate whether ICS may have affected trial results (section 2.3).

### 2.1. The RESTORE trial

The randomised evaluation of sedation titration for respiratory failure (RESTORE) trial was a CRT that compared protocolised sedation with usual care for critically ill children mechanically ventilated for acute respiratory failure [18]. It randomised 31 paediatric intensive care units in the United States, enrolling 2449 individuals (cluster sizes: 12-272). A total of 33 clinical outcomes were analysed (11 continuous, 11 binary, and 11 count or incidence rate outcomes; see Section 3 and Table S1-2 in the Supplemental File for the full list).

### 2.2. Modelling the association between cluster size and outcomes/treatment effects

An association between cluster size and outcome (e.g. if larger clusters have better outcomes on average than smaller clusters) is referred to as “type A” ICS; an association between cluster size and the treatment effects (e.g. if larger clusters respond better to treatment) is referred to as “type B” ICS. Type A ICS can lead to differences for non-collapsible summary measures (e.g. odds ratio), while type B ICS can lead to differences for both collapsible (e.g. difference in means or proportions) or non-collapsible measures.

For each of the 33 outcomes, we fit regression models including cluster size as the sole covariate to evaluate type A ICS or models including cluster size and an additional treatment– cluster size interaction to evaluate type B ICS. No other baseline covariates were adjusted, as ICS is the association between cluster size and outcome/treatment effect unconditional on other covariates. We modelled cluster size using restricted cubic splines [19] with 5 knots to allow for a non-linear association. [20, 21] All models were fitted using IEEs via GEEs with an independence correlation structure, specifying a Gaussian family with an identity link, binomial family with a logit link, and Poisson family with a log link for continuous, binary, and count/incidence outcomes, respectively. The presence of type A or B ICS was tested using a joint Wald test on the spline terms or treatment–spline interaction terms, with a significance threshold of 0.05. The Fay–Graubard bias-corrected sandwich variance estimator was used due the small number of clusters. [22]

The presence of ICS does not necessarily imply a difference between individual- and cluster-average effects. For collapsible measures, type A ICS alone will not cause such differences. Similarly, a non-monotone association (e.g. U-shaped) between cluster size and outcomes/treatment effects could leave the two effects coinciding despite ICS being present. This modelling approach therefore serves as an exploratory tool rather than a definitive test of whether ICS affects results, motivating the separate impact assessment below.

While ICS can lead to differences between individual- and cluster-average treatment effects, the presence of ICS does not necessarily imply a difference between these two effects. For collapsible summary measures, type A ICS alone will not lead to such differences. Another less likely but theoretically possible example is if the association between cluster size and outcomes/treatment effects is non-monotone (e.g. U-shaped, such that small and large clusters have large treatment effects while medium-sized clusters have small effects), it could leave the two effects coinciding despite ICS being present. This modelling approach therefore serves as a tool to explore the potential for ICS and cannot be used to determine whether ICS will lead to differences between the individual- and cluster-average effects, or to what extent these effects might differ. We therefore performed a separate evaluation to determine the impact of ICS on results (next section).

### 2.3. Assessment of the impact of ICS

#### 2.3.1 Comparisons and motivation

For each outcome, we assessed the potential impact of ICS by directly comparing the estimated individual- and cluster-average treatment effects. Since a difference between individual- and cluster-average effects implies ICS, a direct comparison may provide additional evidence of ICS (beyond the results from modelling approach) and demonstrate how results may change under a different estimand.

In addition, we also compared estimates from commonly used but potentially biased methods (GEEs(exch) and mixed-effects models) against unbiased IEE estimates to assess the extent to which the inference from these commonly used estimators was affected by ICS for the individual-average estimand.

#### 2.3.2 Implementation of Estimators

We implemented three different estimators per outcome: (i) unweighted IEEs (unbiased for the individual-average effect), (ii) weighted IEEs (unbiased for the cluster-average effect) and (iii) mixed-effect models for continuous or GEEs(exch) (for binary and count/incidence outcomes [23–25] with the same family and link specification as in section 2.2.

We estimated the difference in means for continuous outcomes, the odds ratio (OR) for binary outcomes, the rate ratio (RR) for count outcomes, and the incidence rate ratio (IRR) for incidence rate outcomes. All target estimands were marginal. We calculated cluster-robust SEs using the Fay–Graubard bias correction for both GEEs(exch) and IEEs, and used the Satterthwaite correction for mixed-effect models. [26] Table 1 summaries the estimators, implementation details, and Stata code.

**Table 1.**
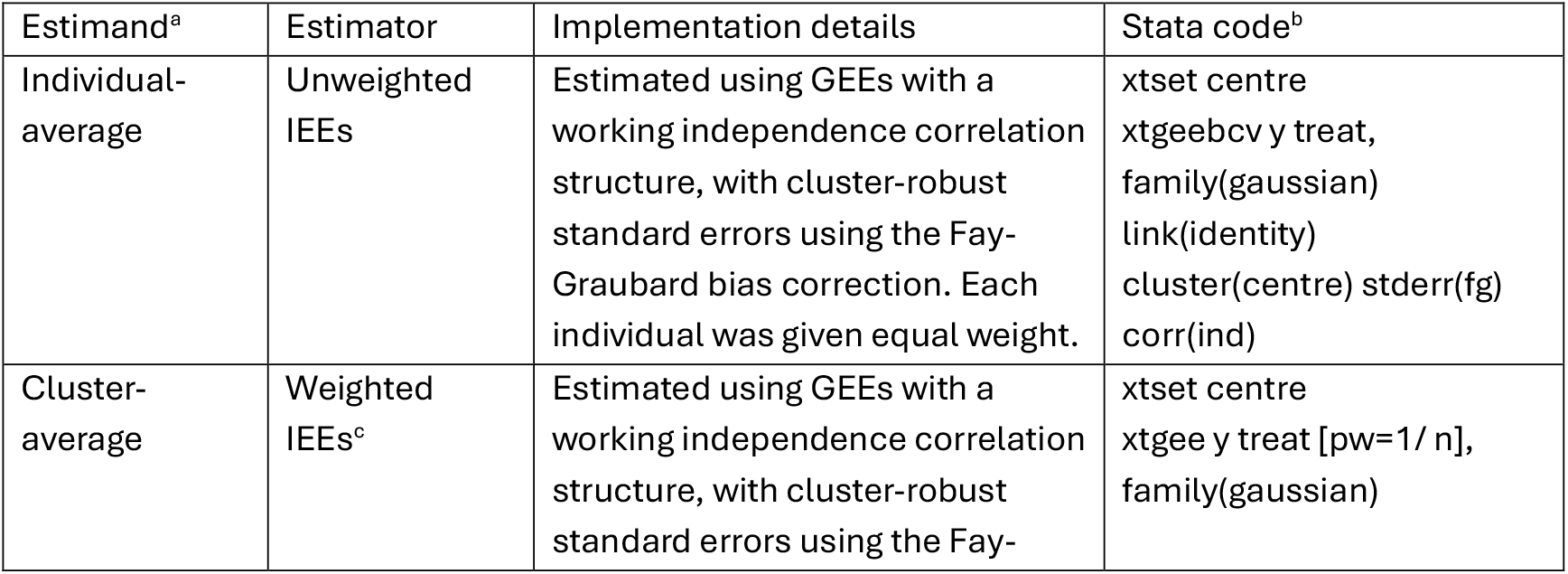
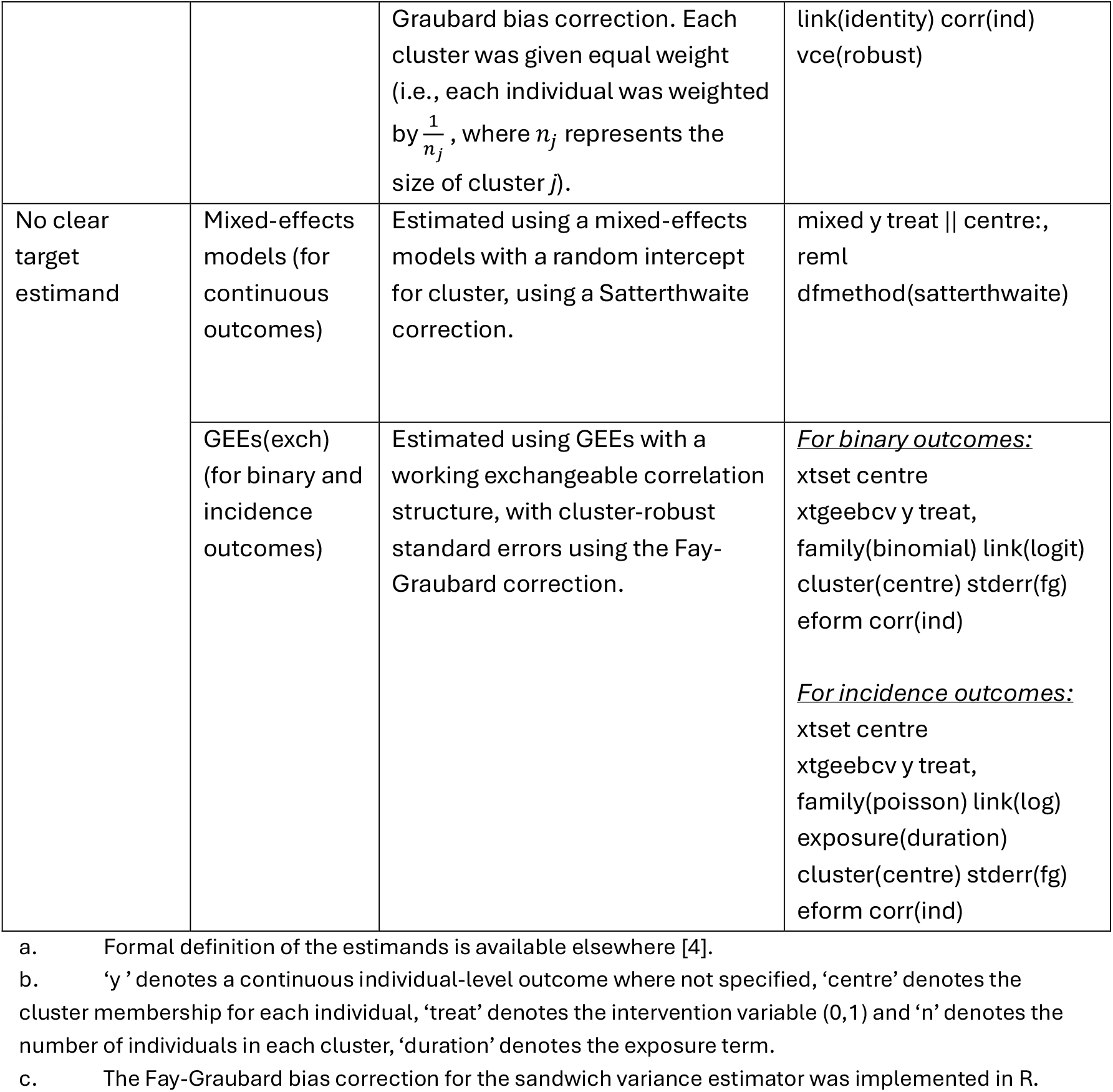
Overview of estimands, estimators and implementation details.

#### 2.3.3 Expression of the difference between estimates

For each comparison (i) weighted vs. unweighted IEE and (ii) GEEs(exch)/mixed-effects vs. unweighted IEE, the unweighted IEE individual-average estimate (IA) served as the reference. The relative percentage difference was calculated as (COMP − IA) / IA × 100%, where COMP represents the comparator estimate. Bias-corrected and accelerated bootstrap CIs were used to assess whether observed differences were attributable to chance, with clusters being resampled 1000 times with replacement; a CI excluding 0 suggests that the difference is unlikely to be due to chance and is likely to have arisen from ICS.

## 3. Results

### 3.1. Binary outcomes

Overall, 4 of 11 outcomes (36%) showed statistically significant evidence of the presence of ICS (3 type A ICS and 1 type B ICS) (Table 2).

**Table 2.** Difference in estimates from individual-average estimators versus cluster-average estimators and GEEs (exch) with bootstrapped CIs for binary outcomes. Treatment effects are presented as odds ratios with 95% CIs.

| Binary | Individual average (unweighted IEE)<sup>a</sup> | Cluster average (weighted IEE)<sup>a</sup> | GEEs (exch)<sup>a</sup> | % Difference (cluster-average vs individual-average) | % Difference (GEE (exch) vs individual-average) | Type A ICS p value<sup>b</sup> | Type B ICS p-value<sup>c</sup> |
| --- | --- | --- | --- | --- | --- | --- | --- |
| Neurotest | 0.72 (0.52, 1.00) | 0.74 (0.52, 1.05) | 0.72 (0.52, 0.99) | 2 (-12, 25) | 0 (-12, 21) | 0.005 | 0.935 |
| Death at 28d | 0.74 (0.43, 1.25) | 0.63 (0.34, 1.18) | 0.72 (0.41, 1.27) | -14 (-38, 19) | -26 (-26, 19) | 0.241 | 0.867 |
| Death at 90d | 0.75 (0.46, 1.23) | 0.60 (0.32, 1.13) | 0.71 (0.41, 1.22) | -19 (-41, 15) | -5 (-31, 16) | 0.502 | 0.438 |
| Inadequate pain management | 1.14 (0.48, 2.70) | 1.22 (0.61, 2.42) | 1.16 (0.56, 2.40) | 7 (-29, 65) | 2 (-41, 67) | 0.450 | 0.602 |
| Inadequate sedation management | 1.30 (0.62, 2.70) | 0.99 (0.54, 1.82) | 1.03 (0.54, 1.95) | -24 (-50, 3) | -21 (-55, 21) | 0.406 | 0.042 |
| Clinically significant iatrogenic withdrawal | 1.35 (0.68, 2.66) | 0.81 (0.38, 1.72) | 0.91 (0.42, 1.97) | -40 (-61, -21) | -32 (-62, 5) | 0.718 | 0.613 |
| Extubation failure | 0.93 (0.64, 1.34) | 0.94 (0.63, 1.40) | 0.91 (0.64, 1.30) | 1 (-19, 34) | -2 (-21, 7) | 0.984 | 0.968 |
| Postextubation stridor | 1.65 (1.02, 2.67) | 1.38 (0.87, 2.19) | 1.57 (0.98, 2.50) | -16 (-38, 7) | -5 (-28, 12) | 0.944 | 0.343 |
| Immobility-related stage 2 or worse pressure ulcers | 0.26 (0.10, 0.70) | 0.11 (0.03, 0.36) | 0.21 (0.07, 0.57) | -58 (-81, -33) | -21 (-58, 22) | <0.001 | 0.380 |
| New tracheostomy | 0.48 (0.15, 1.50) | 0.42 (0.15, 1.25) | 0.43 (0.14, 1.35) | -10 (-48, 38) | -9 (-49, 47) | 0.001 | 1.000 |
| iatrogenic withdrawal syndrome | 1.02 (0.66, 1.59) | 1.13 (0.72, 1.77) | 1.04 (0.68, 1.59) | 11 (-12, 42) | 2 (-19, 27) | 0.601 | 0.051 |

CI: confidence interval; IEE: independent estimating equations; ICS: informative cluster size.
<sup>a</sup> CIs formulated using standard errors with the Fay-Graubard small sample correction.
<sup>b</sup> Obtained from joint tests on the restricted cubic spline terms of cluster size.
<sup>c</sup> Obtained from joint tests on the interaction between treatment and spline terms of cluster size.

Figure 1 shows the association between cluster size and (a) outcome and (b) treatment effect, for “immobility-related stage 2/worse pressure ulcers”: type A ICS was statistically significant (P<0.001), there was no clear evidence of type B ICS (P=0.38) despite some association between cluster size and treatment effect.

**Figure 1.**
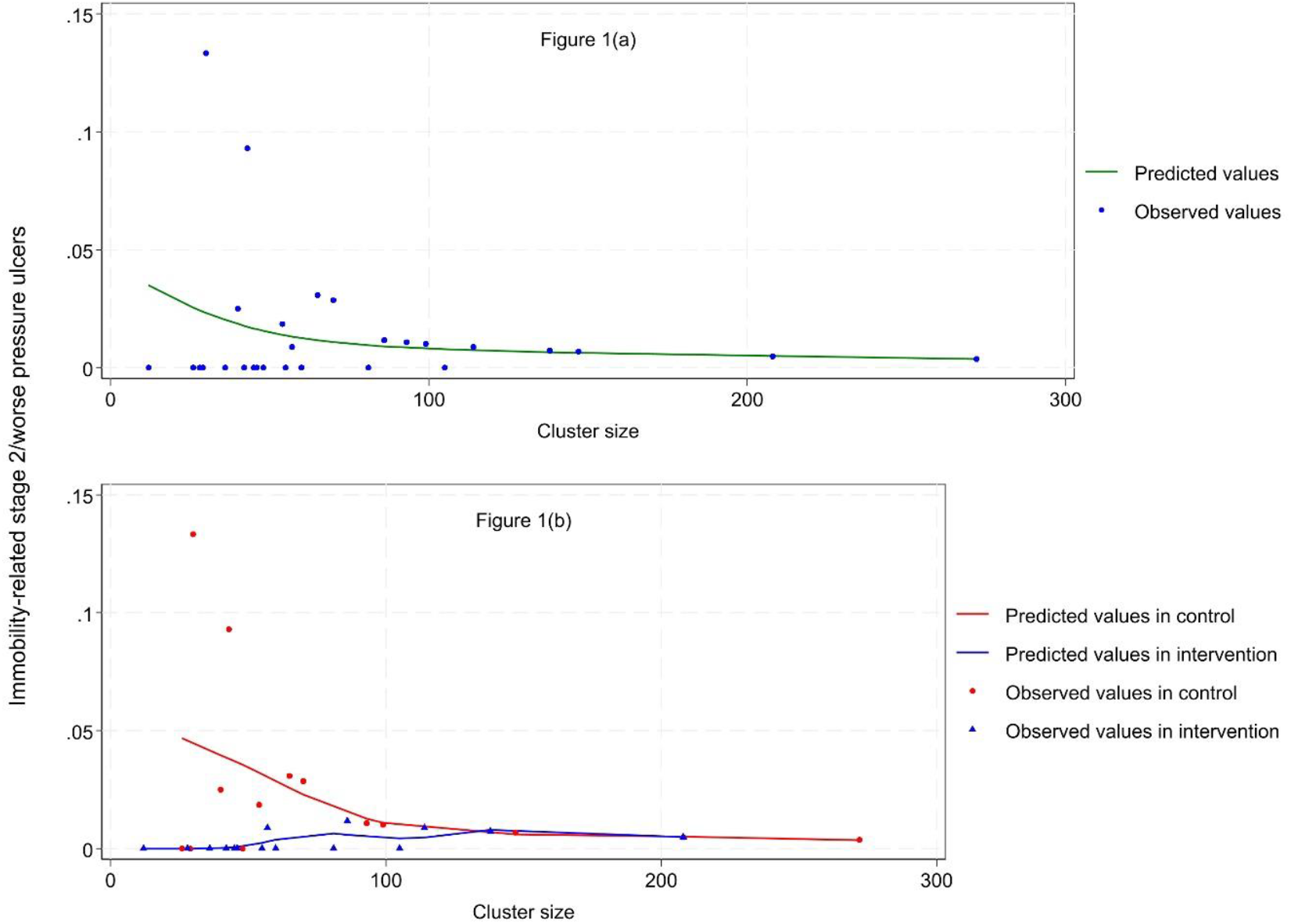
Association between cluster size and (a) outcomes and (b) treatment effect, for the outcome immobility-related stage 2/worse pressure ulcers. To evaluate the association between cluster size and the outcome, we pooled across both treatment arms together in (a).

There was statistically significant evidence of a difference between the individual- and cluster-average estimands for 2 of the 11 binary outcomes (18%) (Figure 2 and Table 2), both with differences >40% (clinically significant iatrogenic withdrawal and immobility-related stage 2/worse pressure ulcers). Overall, 8 outcomes (73%) showed a relative difference >10% between these two estimands. No outcomes showed a statistically significant difference between the GEEs(exch) estimate and the individual-average estimate (from IEEs), though 4 outcomes (36%) showed a difference >10% (Table 2).

**Figure 2.**
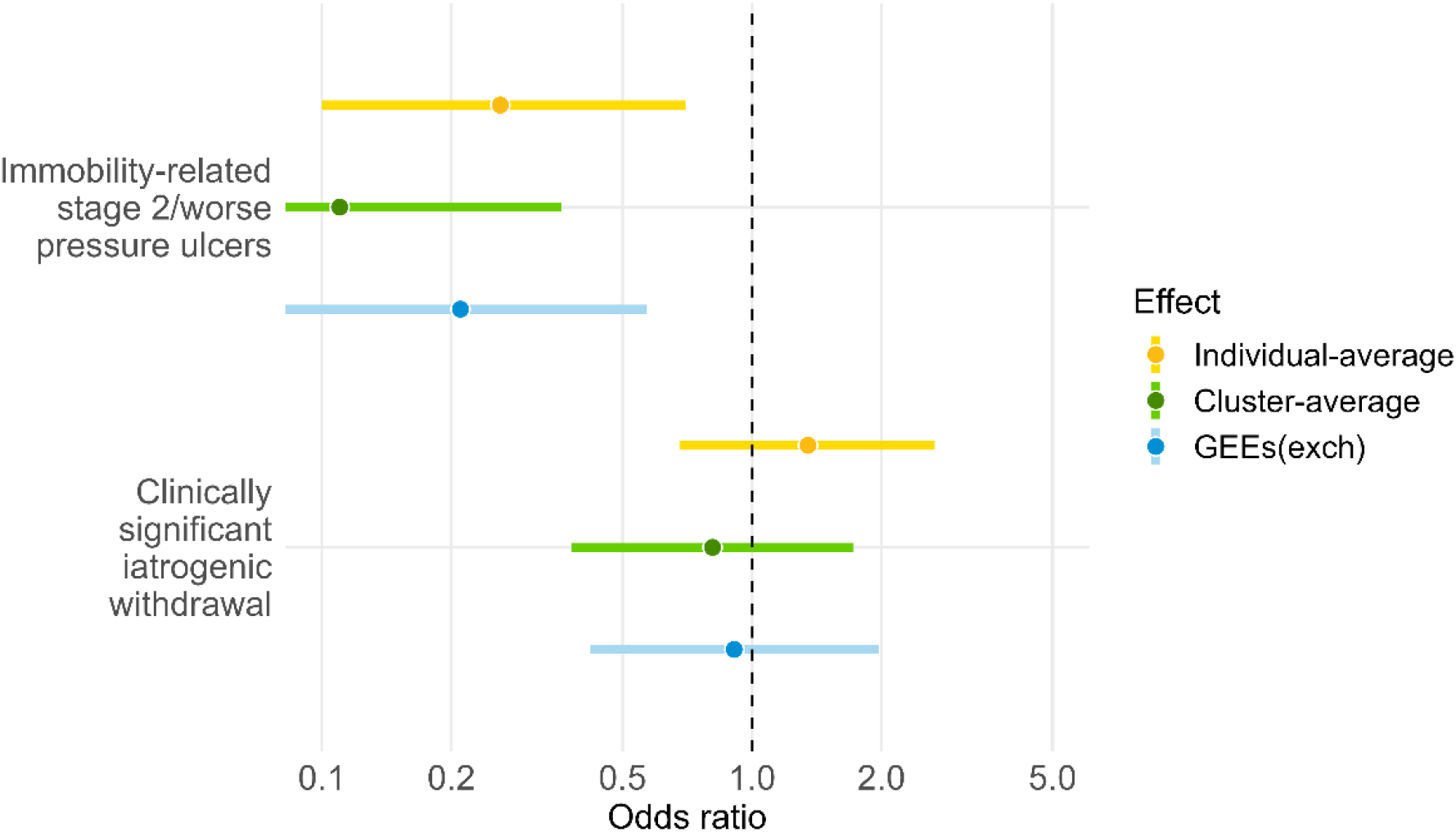
Treatment effect estimates with 95% CIs for binary outcomes with statistically significant evidence of a difference between individual- and cluster-average estimand.

For clinically significant iatrogenic withdrawal, the individual- and cluster-average OR estimates were in opposite directions (individual-average OR 1.35, 95% 0.68 to 2.66 vs. cluster-average OR 0.81, 95% CI 0.38 to 1.72). The GEEs(exch) OR was in between the two (OR 0.91, 95% CI 0.42 to 1.97).

For immobility-related stage 2/worse pressure ulcers, the individual-average OR was 0.26 (95% CI 0.10 to 0.70), whereas the cluster-average and GEEs(exch) ORs were 0.11 (95% CI 0.03 to 0.36) and 0.21 (95% CI 0.07 to 0.57), respectively, denoting percentage differences of 58% and 21%.

Notably, for postextubation stridor, although there was no clear evidence of ICS, the choices of estimand and/or estimator affected interpretations, changing the statistical significance of the intervention effect. The individual-average OR was 1.65 (95% CI 1.02 to 2.67), demonstrating statistically significant harm from the intervention, whereas the cluster-average OR (1.38, 95% CI 0.87 to 2.19) and GEEs(exch) OR (1.57, 95% CI 0.98 to 2.50) were both closer to the null and not statistically significant.

### 3.2. Continuous outcomes

Overall, 8 of 11 outcomes (73%) showed statistically significant evidence ICS (7 type A ICS and 1 type B) (Table S1 in the Supplemental file). There was statistically significant evidence of a difference between the individual- and cluster-average estimands for 3 of 11 continuous outcomes (27%) (Figure 3 and Table S1 in the Supplemental file), two of which showed>150% differences, partly because small individual-average effects serve as the denominator. Overall, 6 outcomes (55%) showed a relative difference >10% between the individual- and cluster-average estimands. There was statistically significant evidence of a difference between mixed-effects model estimate and the individual-average estimate for 2 of 11 outcomes (18%), with 6 outcomes (55%) showing a relative difference >10% (Table S1 in the Supplemental File).

**Figure 3.**
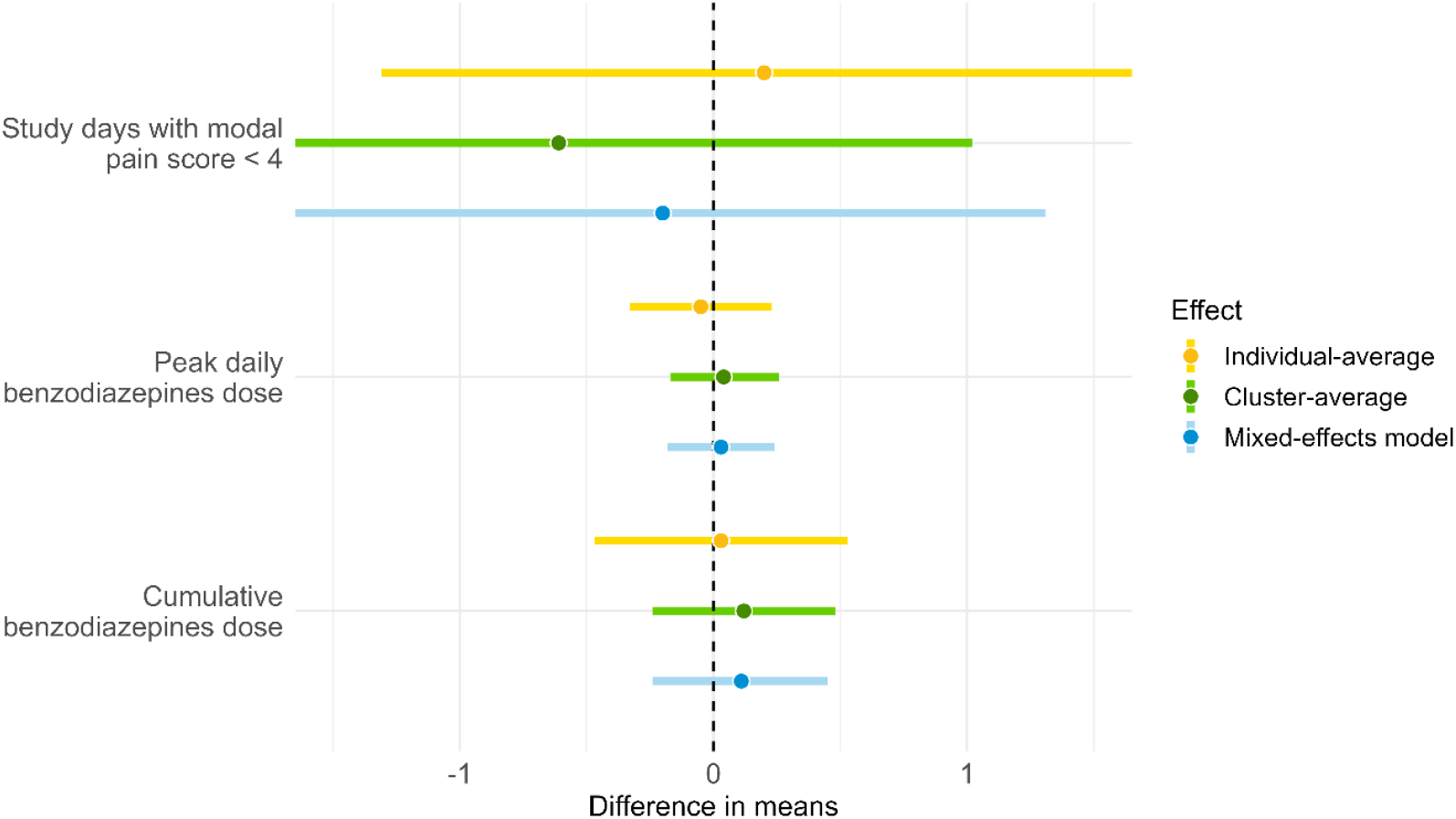
Treatment effect estimates with 95% CIs for continuous outcomes with statistically significant evidence of a difference between individual- and cluster-average estimands.

For peak daily benzodiazepines dose, the individual-average and cluster-average estimates were in opposite directions (individual-average −0.05, 95% CI −0.33 to 0.23 vs. cluster-average 0.04, 95% CI −0.17 to 0.26). The mixed-effect model estimate was between the two (0.03, 95% CI −0.18 to 0.24). For cumulative benzodiazepines dose, the difference between individual- and cluster-average effects exceeded 300 % (individual-average 0.03, 95% CI −0.47 to 0.53 vs. cluster-average 0.12, 95% CI −0.24 to 0.48). The mixed-effect model estimate was between the two (0.11, 95% CI −0.24 to 0.45).

### 3.3. Count and incidence rate outcomes

Overall, 4 of 11 outcomes (36%) showed statistically significant evidence of informative cluster (3 type A ICS, 1 showing both type A and B ICS) (Table S2 in the Supplemental file).

No outcomes showed statistically significant differences between the individual- and cluster-average estimands, though 4 outcomes (36%) showed a relative difference >10%. Similarly, no outcomes showed statistically significant difference between the GEEs(exch) estimate and individual-average estimate (from IEEs), with 3 outcomes (18%) showing a difference >10% (Table S2 in the Supplemental File).

Although no outcomes showed statistically significant differences between estimands, large differences between estimators were observed for some outcomes. For the outcome number of catheter-associated bloodstream infection, the individual-average RR was 1.12 (95% CI 0.20 to 6.32) and cluster-average RR was 2.19 (95% CI 0.45 to 10.80), denoting a percentage difference of 96%; the GEEs(exch) RR was 1.43 (95% CI 0.28 to 7.37), denoting a 28% percentage difference to the individual-average RR.

## 4. Discussion

Different estimands can be targeted in CRTs, including the individual- and cluster-average estimands. When ICS is present, the individual- and cluster-average estimands can differ in magnitude, and commonly used treatment coefficient estimators such as mixed-effects models and GEEs(exch) can be biased for both estimands. With few published studies formally examining ICS, its prevalence remains unknown. In this re-analysis of the RESTORE trial, we assessed whether ICS was present and investigated its impact on trial results.

We found evidence of an association between cluster size and either outcomes or treatment effects for 16 of 33 outcomes (48%). This led to statistically significant differences between the individual- and cluster-average estimands for 5 of 33 outcomes (15%), and differences >10% for 17 outcomes (52%). We also identified differences between estimates from methods that can be affected by ICS (mixed-effects model or GEEs(exch)) and estimates from unweighted IEEs (which are unaffected by ICS). Overall, we found statistically significant evidence that mixed-effects models/GEEs(ecxh) were biased for 2 outcomes (6%) relative to the individual-average effect (unweighted IEE), with notable differences (>10%) for 13 outcomes (39%).

In some cases, the choice of estimand or estimator affected the interpretation of results, either by changing whether results were statistically significant or switching the direction of the estimated effect. For the adverse event outcome postextubation stridor, the individual-average estimate showed statistically significant evidence of harm, whereas the cluster-average and GEEs(exch) estimates did not. For clinically significant iatrogenic withdrawal, the individual-average OR was 1.35 (95% CI 0.68–2.66), in the opposite direction to the cluster-average OR (0.81, 95% CI 0.38–1.72) and GEEs(exch) OR (0.90, 95% CI 0.42–1.97).

These findings highlight the importance of carefully considering both the estimand and the estimator in CRTs. CRTs investigators should clearly define their target estimand from the outset of the study and choose an estimator aligned with that estimand, which involves assessing the plausibility of the assumptions underpinning the estimator, such as the likelihood of ICSs. In addition to the individual- and cluster-average treatment effects, other aspects of defining estimands in CRTs have been recognised, such as the population of clusters and the marginal vs cluster-specific treatment effect [4, 27, 28]; these should also be considered to ensure that the estimands are unambiguous and relevant to the trial’s research question.

This study had several limitations. The analysis is likely to be given the small number of clusters (n=31). Despite this, it found 5 of 33 outcomes (15%) with statistically significant evidence that results were affected by ICS based on a significant difference between individual- and cluster-average estimands (at 0.05 significance level). It also provides a template for those who plan to perform secondary analysis to investigate ICS in completed CRTs. Notably, ICS can potentially be introduced by missing data. For instance, some clusters might be smaller because individuals with poor outcomes were missing from them, resulting in smaller (observed) clusters with better outcomes than the others. However, missing data was minimal in our analysis (e.g., there was no missingness on outcomes for which we found statistically significant difference between the individual- and cluster-average estimands), indicating that ICS was unlikely due to missing data.

Another potential limitation is the apparent discrepancies observed for some outcomes between the two ICS evaluation approaches (i.e. modelling the association between cluster size and outcomes/treatment effects vs. direct comparison of individual- and cluster-average effects). For clinically significant iatrogentic withdrawal, we found a statistically significant difference between the individual- and cluster-average estimates, but no clear association between cluster size and outcomes. This may be due in part to differences in power between the two methods, as well as differences in what each method evaluates: a direct comparison of treatment effect estimates assesses both the presence of ICS and impact of ICS on results for a given outcome, while modelling the cluster size-outcomes/treatment effects association only tests for the presence of ICS.

Our findings suggest several areas for future research. First, further empirical evaluations of CRTs are needed to examine the prevalence of ICS and its impact on trial results. Second, studies are needed to evaluate the efficiency of ICS-robust methods (IEEs, cluster-level summary analyses, and novel methods such as model-robust standardisation [29]) relative to mixed-effects models and GEEs(exch); if efficiency losses are modest, it may be worth switching to the robust methods given our findings. Simulations would be needed to appropriately compare precision across methods. Third, comparing SEs across estimators can be challenging when different small-sample corrections are applied; the Fay–Graubard correction inflates SEs, whereas the Satterthwaite correction modifies degrees of freedom, affecting CIs and p-values differently. Evaluation of SE correction methods would help identify optimal approaches.

## 5. Conclusion

In this re-analysis, we found empirical evidence that certain outcomes were affected by ICS, leading to differences in results based on the choice of estimand and estimator. This suggests the need to clearly specify the target estimand and carefully consider the likelihood of ICSs when choosing an estimator in cluster-randomised trials.

## Data Availability

Data from the RESTORE trial was obtained from the U.S. National Heart, Lung and Blood Institute, Biologic Specimen and Data Repository Information Coordinating Center (BioLINCC) (https://biolincc.nhlbi.nih.gov/home/). The authors do not have permission to share this data directly; however, the data are available to researchers upon request and approval from BioLINCC.

## CRediT authorship contribution statement

Dongquan Bi: Writing – original draft, Writing – review & editing, Data curation, Formal analysis, Methodology.

Brennan Kahan: Writing – review & editing, Conceptualization, Supervision, Methodology.

Andrew Copas: Writing – review & editing, Conceptualization, Supervision, Methodology.

Fan Li: Writing – review & editing, Methodology.

Michael Harhay: Writing – review & editing, Data curation.

