## Supplemental File for "Choice of estimands and estimators affected the interpretation of results for some outcomes in a cluster-randomised trial (RESTORE) due to informative cluster size"

### **Table S1. Difference in estimates from individual-average estimators versus cluster-average estimators and mixed-effects models with bootstrapped CIs for continuous outcomes. Treatment effects are presented as differences in means with 95% CIs.**

| **Continuous outcomes (SD)** | **Individual-average (unweighted IEE)^a^** | **Cluster-average (weighted IEEs)^a^** | **Mixed-effect models^b^** | **% Difference (cluster-average vs individual-average)** | **% Difference (mixed model vs individual-average)** | **Type A ICS P value^c^** | **Type B ICS P-value^d^** |
| --- | --- | --- | --- | --- | --- | --- | --- |
| Mean daily opioids dose  (0.59) | -0.08 (-0.33, 0.17) | -0.12 (-0.27, 0.03) | -0.11 (-0.27, 0.05) | 48 (-118, 4992) | 34 (-145, 5607) | 0.014 | 0.994 |
| Peak daily opioids dose  (0.77) | -0.11 (-0.43, 0.22) | -0.15 (-0.36, 0.06) | -0.14 (-0.35, 0.07) | 44 (-128, 4376) | 31 (-132, 5200) | 0.039 | 0.998 |
| Cumulative opioids dose  (1.37) | -0.19 (-0.73, 0.35) | -0.28 (-0.60, 0.04) | -0.26 (-0.59, 0.08) | 52 (-171, 5469) | 38 (-164, 5457) | 0.004 | 0.96 |
| Mean daily benzodiazepines dose  (0.59) | 0.04 (-0.17, 0.26) | 0.10 (-0.06, 0.26) | 0.09 (-0.06, 0.25) | 143 (-10, 4630) | 125 (-9.4, 24595) | 0.03 | 0.855 |
| Peak daily benzodiazepines dose  (0.78) | -0.05 (-0.33, 0.23) | 0.04 (-0.17, 0.26) | 0.03 (-0.18, 0.24) | -188 (-71645, -53) | -162 (-8841, -60) | 0.002 | 0.878 |
| Cumulative benzodiazepines dose  (1.38) | 0.03 (-0.47, 0.53) | 0.12 (-0.24, 0.48) | 0.11(-0.24, 0.45) | 301 (118, 12102) | 253 (81, 54405) | 0.044 | 0.978 |
| Peak WAT-1 score (0.55) | 0.00 (-0.13, 0.13) | 0.02 (-0.11, 0.15) | 0.01 (-0.12, 0.13) | NA | NA | 0.668 | 0.174 |
| Study days awake and calm ^a^  (0.29) | 11.61 (3.75, 19.46) | 9.83 (2.49,17.20) | 9.89 (2.60, 17.17) | -2 (-6, 2) | 2 (-7, 3) | <0.001 | 0.692 |
| Study days with modal pain score <4 ^d^  (0.11) | 0.20 (-1.31, 1.70) | -0.61(-2.24, 1.02) | -0.20 (-1.72, 1.31) | -1 (-2, -0.01) | 0 (-1, 0.4) | 0.909 | 0.616 |
| Study days with an episode of pain ^d^  (0.28) | 18.54 (10.79, 26.28) | 18.29 (9.07, 27.50) | 17.91 (8.68, 27.15) | 0 (-5, 5) | -1 (-6, 5) | 0.957 | 0.154 |
| Study days with an episode of agitation ^d^  (0.32) | 14.00 (3.90, 24.11) | 13.06 (3.94, 22.20) | 13.02 (3.80, 22.24 | -1 (-7, 4) | -1 (-8,5) | 0.990 | 0.154 |
| Study days with WAT-1 score ≥3 ^e^  (0.29) | -0.52 (-8.96, 7.91) | -0.12 (-7.15, 6.92) | -0.84(-8.35, 6.67) | 0 (-4, 4) | 0 (-7,3) | 0.224 | 0.005 |

SD: standard deviation; CI: confidence interval; IEE: independent estimating equations; ICS: informative cluster size; NA: models did not converge

^a^ CIs formulated using standard errors with the Fay-Graubard small sample correction.

^b^ CIs formulated using standard errors with Satterthwaite small sample correction.

^c^ Obtained from joint tests on the restricted cubic spline terms of cluster size.

^d^ Obtained from joint tests on the interaction between treatment and spline terms of cluster size

^e^ The direct difference instead of percentage difference was reported for the outcome. This was done as the outcome was a percentage outcome, with values ranging from 0 to 100 (e.g., percentage of days with modal pain score <4), making the percentage difference hard to interpret.

### **Table S2. Difference in estimates from Individual-average estimators versus cluster-average estimators and GEEs (exch) for count and incidence rate outcomes. Treatment effects are presented as rate ratios for count outcomes and incidence rate ratios for incidence rate outcomes with 95% CIs.**

| **Count or incidence outcome** | **Individual average (unweighted IEE)^a^** | **Cluster average (weighted IEE)^a^** | **GEEs (exch)** | **% Difference (cluster-average vs individual-average)** | **% Difference (GEE (exch) vs individual-average)** | **Type A ICS P value^b^** | **Type B ICS P-value^c^** |
| --- | --- | --- | --- | --- | --- | --- | --- |
| No. unplanned endotracheal tube extubations | 0.75 (0.35, 1.61) | 0.86 (0.46, 1.62) | 0.96 (0.50, 1.82) | 14 (-24, 5) | 27(-19, 2034) | 0.852 | 0.269 |
| No. unplanned removals of any invasive tube | 1.36 (0.68, 2.71) | 1.65 (0.67, 4.11) | 1.53 (0.70, 3.31) | 21(-36, 103) | 12(-42, 99) | 0.278 | 0.208 |
| No. ventilator-associated pneumonias | 0.73 (0.22, 2.42) | 0.56 (0.19, 1.69) | 0.70 (0.22, 2.30) | -23 (-61, 52) | -4(-38, 31) | 0.130 | 0.535^d^ |
| No. catheter-associated bloodstream infections | 1.12 (0.20, 6.32) | 2.19 (0.45,10.80) | 1.43 (0.28, 7.37) | 96 (-20, 715) | 28(-24, 149) | 0.001 | 0.049^d^ |
| Time to successfully extubated within 28 days | 1.07 (0.85, 1.33) | 1.15 (0.96, 1.39) | NA | 8 ( -3, 17) | NA | 0.093 | 0.958 |
| Time to recover from acute respiratory failure within 28 days | 1.01 (0.74, 1.37) | 1.07 (0.79, 1.44) | NA | 6 (-4, 23) | NA | 0.063 | 0.986 |
| Time to wean from mechanical ventilation within 28 days | 1.11 (0.89, 1.39) | 1.09 (0.95, 1.40) | NA | -2 (-18, 12) | NA | 0.446 | 0.467 |
| Stayed in pediatric ICU less than 90 days | 1.07 (0.86, 1.33) | 1.08 (0.88, 1.33) | NA | 1(-11, 13) | NA | 0.001 | 0.923 |
| Stayed in hospital less than 90 days | 1.13 (0.91, 1.41) | 1.10 (0.90, 1.34) | NA | -3 (-16, 7) | NA | 0.017 | 0.466 |
| Stopped opioid by 28 days | 1.28 (1.04, 1.57) | 1.32 (1.11, 1.58) | NA | 4 (-7, 14) | NA | 0.005 | 0.908 |
| Awake/calm state by 28 days | 1.20 (0.87, 1.67) | 1.24 (0.92, 1.67) | NA | 3 (-14, 17) | NA | 0.892 | 0.737 |

SD: standard deviation; CI: confidence interval; IEE: independent estimating equations; ICS: informative cluster size; NA: models did not converge

^a^ CIs formulated using standard errors with the Fay-Graubard small sample correction.

^b^ Obtained from joint tests on the restricted cubic spline terms of cluster size.

^c^ Obtained from joint tests on the interaction between treatment and spline terms of cluster size

^d^ Number of knots were reduced from 5 to 3 for restricted cubic splines of cluster size due to convergence issues.
